# Diarrhea may be underestimated: a missing link in 2019 novel coronavirus

**DOI:** 10.1101/2020.02.03.20020289

**Authors:** Weicheng Liang, Zhijie Feng, Shitao Rao, Cuicui Xiao, Ze-Xiao Lin, Qi Zhang, Qi Wei

**Affiliations:** Vaccine Research Institute, The 3rd Affiliated Hospital of Sun Yat-sen University, Sun Yat-sen University, Guangzhou, China; Cell-gene Therapy Translational Medicine Research Center, The 3rd Affiliated Hospital of Sun Yat-sen University, Guangzhou, China; Guangdong Provincial Key Laboratory of Liver Disease Research, Guangzhou, China; School of Biomedical Sciences, The Chinese University of Hong Kong, Shatin, Hong Kong SAR, China; Department of Medical Oncology, The 3rd Affiliated Hospital of Sun Yat-sen University, Sun Yat-sen University, Guangzhou, China; Department of Gastroenterology, The Second Hospital of Hebei Medical University; Hebei Key Laboratory of Gastroenterology, Hebei Institute of Gastroenterology, Shijiazhuang, China

## Abstract

The outbreak of pneumonia caused by the 2019 Novel Coronavirus (2019-nCoV) was reported in Wuhan City, China. However, the clinical symptoms varied in different reports. Based on results of inter-group difference test, we found that the incidence of diarrhea differed in three recent reports. As 2019-nCoV utilizes the same cell entry receptor ACE2 as severe acute respiratory syndrome coronavirus (SARS-CoV) and ACE2 tightly controls intestinal inflammation, to trace the route of infection mediated by 2019-nCoV, we used the single-cell RNA sequencing data for analysis. We found that the ACE2 mRNA was highly expressed in the healthy human small intestine rather than the lung. Besides, single-cell RNA sequencing data showed that ACE2 was significantly elevated in the proximal and distal enterocytes, where the small intestinal epithelium is exposed to the foreign pathogen. Thus, we suspect that ACE2-expressing small intestinal epithelium cells might be vulnerable to 2019-nCoV infection when people eat infected wild animals and diarrhea may serve as an indicator for infection, suggesting that clinicians should pay more attention to patients with diarrhea during the outbreak of pneumonia.

## Introduction

In December 2019, the first pneumonia case caused by a previously unknown coronavirus 2019-nCoV occurred in Wuhan City, China. Soon afterward, a series of pneumonia cases caused by 2019-nCoV emerged in other provinces in mainland China. Subsequently, individuals who were infected by 2019-nCoV traveled globally, and a series of exported infection cases have been identified in Japan, Korea, the United States, Canada, France, etc. On January 30, 2020, the WHO has confirmed 7,818 cases worldwide, and 7,736 cases were reported in China.

According to clinical observation, the most common symptoms were fever, cough, and myalgia or fatigue. Less common symptoms included sputum production, headache, hemoptysis, and diarrhea. Based on three recent publications^1-3^, we compared the incidence of these common symptoms and found that the incidence of diarrhea significantly differed in these three reports. As some patients suffering from diarrhea will first seek help from the gastroenterologist, incorrect estimation of diarrhea may increase the risk of 2019-nCoV infection in the gastroenterology department. Therefore, a comprehensive understanding of the clinical features of pneumonia patients will further improve hospital infection control and reduce healthcare-associated infection.

By using the next-generation sequencing technology, scientists revealed that the sequence and genome structure of 2019-nCoV is highly homologous to that of SARS-CoV ^4,5^. It has been reported that Angiotensin-converting enzyme 2 (ACE2) serves as a receptor for SARS-CoV by interacting with the spike protein of SARS-CoV^6^. Because of the high sequence homology, it is likely that the spike protein of 2019-nCoV shares the same cell entry receptor ACE2 with that of SARS-CoV. Consistently, four independent groups have recently provided evidence to support this hypothesis^7-10^.

In the current study, we found that the incidence of diarrhea differed significantly in three recent reports. As 2019-nCoV receptor ACE2 plays a role in intestinal inflammation and diarrhea, we subsequently evaluated the expression profiles of ACE2 by analyzing the RNA-sequencing and single-cell-sequencing data. We found that ACE2 was highly expressed in the small intestine, especially in the proximal and distal enterocytes. To sum up, we proposed that ACE2-expressing small intestinal epithelium cells might be are more vulnerable to attack by 2019-nCoV and clinicians should be careful when their patients complain of diarrhea since the incidence of diarrhea may be underestimated.

## Methods

### Inter-group difference test

The original information of patients infected by 2019-nCoV infection was retrieved from three recent publications^1-3^. The 2019-nCoV infection was laboratory-confirmed by real-time RT-PCR and next-generation sequencing. Categorical variables were expressed as number with percentage in parenthesis. All the variables were compared by Fisher’s exact test for the three studies as one study has a relatively small sample size^1^. Continuous variables were expressed as median with inter-quartile range (IQR) in parenthesis. A two-sided α of less than 0.05 was considered statistically significant. Statistical analyses were performed using the R software, version 3.5.3.

### ACE2 expression profile and

The expression profiles of ACE2 from normal human tissues were obtained from public database NCBI (https://www.ncbi.nlm.nih.gov/). The original expression data was collected and then plotted by GraphPad Prism 5.

### Single-cell sequencing analysis

The public single-cell RNA-seq sequencing data (GSE92332) was downloaded from the GEO database. The 10X matrix file of GSE92332_atlas_UMIcounts.txt was used for subsequent analysis based on R package Seurat (Version 3.1.2). In order to filter out low-quality cells and low-quality genes, strict parameters, “min.featrue=1000” and “min.cell=20”, were used in the function CreateSeuratObject. The data was subsequently log-normalized by the function NormalizeData with the default parameters. The genes with highly variable expression were identified by the function FindVariableGenes. After the data were processed by the function ScaleData, PCA dimensionality reduction was performed utilizing the function RunPCA. Based on the analysis by the function of JackStraw and ScoreJackStraw, the first 15 PCA components were selected for further two-dimensional t-distributed stochastic neighbor embedding (tSNE). The setting of k.param was 30 in the function FindNeighbors, and the setting of the resolution was 1.5 in the function FindClusters. In the downloaded original gene-barcode matrix, Adam L.Haber and his coworkers had annotated the cell identities of each barcode. In our analysis, the cell types were identified by the expression of marker genes and the annotation by the original gene-barcode matrix. Gene expression of different cell types was illustrated by the functions of FeaturePlot and VlnPlot.

## Results

### Different incidence of diarrhea during clinical observation

Based on the recent publications, the most common symptoms in patients infected by 2019-nCoV were fever, cough, myalgia, and fatigue. A few infected people have symptoms like diarrhea and headache. However, the incidence of clinical features differs in different reports. As the clinical symptoms may affect the preliminary diagnostic accuracy, in order to establish accurate preliminary diagnostic criteria, we compared the incidence of these common symptoms. We first collected the data from three recent publications and compared the incidence^1-3^. Interestingly, we found that the incidence of leucopenia, fever, and diarrhea in the three studies showed a statistically significant difference (Table 1). Among these symptoms, diarrhea displayed the smallest p-value (p value= 0.016), suggesting that the criteria for diagnosing diarrhea may differ in different hospitals. Due to the different criteria, clinicians may underestimate the value of this symptom in clinical practice, and it may affect the preliminary diagnostic accuracy.

**Table 1.** Inter-group comparison between three recent publications

| Characteristics | Huang *et al.*<br>(41 cases) | Chan *et al.*<br>(6 cases) | Chen *et al.*<br>(99 cases) | Difference<br>between groups (p value) |
| --- | --- | --- | --- | --- |
| Age | 49.0(17.0) | 46.2(28.3) | 55.5(17.7) | -- |
| Sex(female) | 11/41(27%) | 3(50%) | 32(32%) | 0.4591 |
| Comorbidity | 13/41(32%) | 4(67%) | 50(51%) | 0.0818 |
| Fever | 40/41(98%) | 5(83%) | 82(83%) | *0.0345<sup>a</sup>* |
| Cough | 31/41(76%) | 4(67%) | 81(82%) | 0.3829 |
| Diarrhoea | 1/38(3%) | 2(33%) | 2(2%) | *0.016<sup>b</sup>* |
| Shortness of breath/<br>difficulty in breath | 22/40(55%) | NA | 31(31%) | -- |
| Haemoptysis | 2/39(5%) | NA | NA | -- |
| Sputum production | 11/39(28%) | 2(33%) | NA | -- |
| Myalgia or fatigue | 18/41(44%) | 3(50%)<sup>d</sup> | NA | -- |
| Headache | 3/38(8%) | NA | 8(8%) | -- |
| Leucopenia | 10/40(25%) | 0(0%) | 9(9%) | *0.0443<sup>c</sup>* |
| Platelet count | 2/40(5%) | 0(0%) | NA | -- |

Note: Leucopenia: less white blood cell count ( $<4 \times 10^9/L$ ); Platelet count: $<100 \times 10^9/L$ ;
a: Huang vs. Chan: 0.2414; Chan vs. Chen: 1.00;Huang vs. Chen:0.0235;
b: Huang vs. Chan: 0.044; Chan vs. Chen: 0.016; Huang vs.Chen:1.00;
c: Huang vs. Chan: 0.3145; Chan vs. Chen: 1; Huang vs.Chen:0.026;
d: Symptom: generalized weakness;
NA: not available;
Difference between groups was calculated by Fisher's exact test in R software.

### The expression profiles of ACE2 in healthy people

By using the next-generation sequencing technology, scientists successfully identified and characterized a novel RNA coronavirus with around 29.8 kilobases in length^5^. Phylogenetic analysis of the viral genome indicates that the genome organization of this novel coronavirus is most closely related to that of the bat SARS-CoV. Interestingly, subsequent results showed that 2019-nCoV shared the same cellular entry receptor ACE2 as SARS-CoV^7^.

In terms of the importance of ACE2 in modulating intestinal inflammation and diarrhea^11^, we examined the expression profiles of ACE2 in various human tissues. According to the RNA-sequencing data, we found that ACE2 was highly expressed in the human small intestine (Figure 1). Intriguingly, the RNA level of ACE2 was quite low in lung tissues.

**Figure 1.**
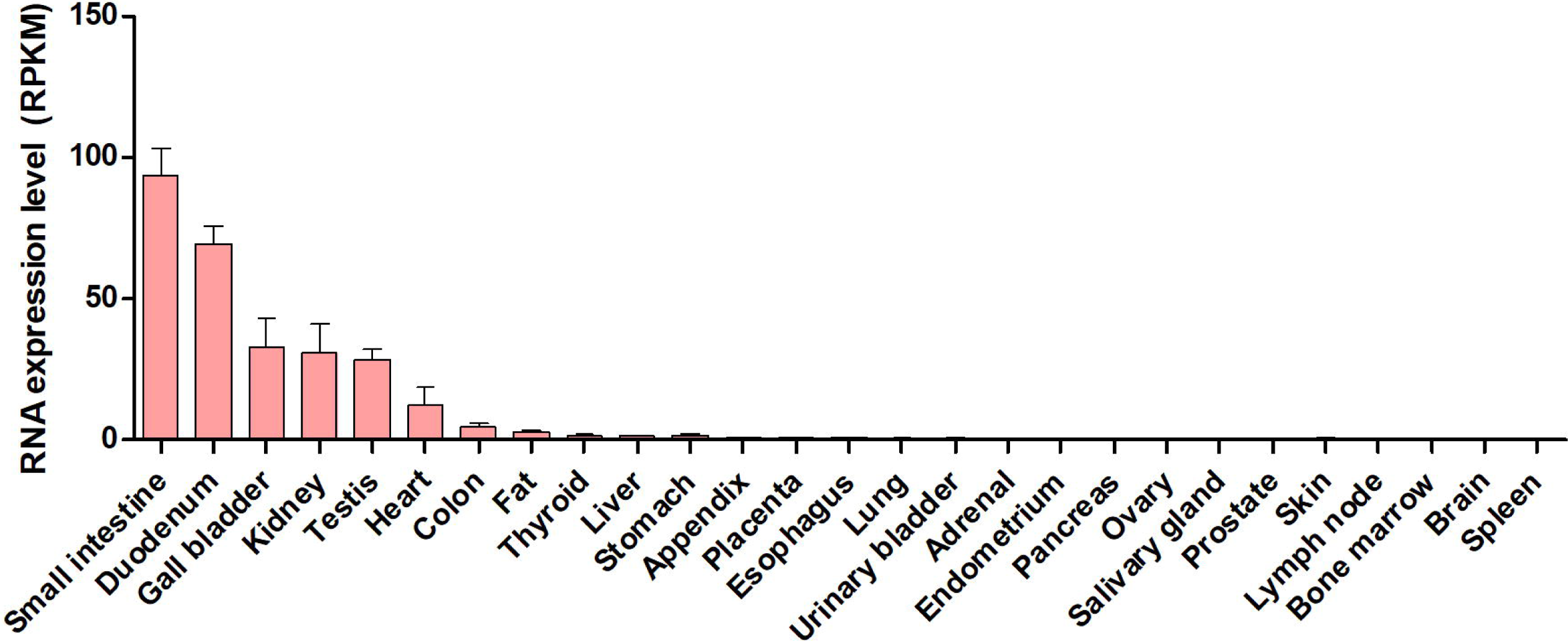
The expression profile of ACE2 in normal human tissues.

### Identification of ACE2-expressing cells in small intestine by single-cell sequencing data

Given that the distribution of ACE2 receptor may determine the route of 2019-nCoV infection and ACE2 is highly expressed in the small intestine, we next evaluated the expression profiles of ACE2 in different cell populations of the small intestine. Currently, single-cell RNA sequencing (scRNA-Seq) technology is a powerful tool that enables us to investigate the ACE2 expression in various cell types and provide quantitative information at single-cell resolution. Based on the scRNA-Seq data (GSE92332)^12^, we utilized the bioinformatics tools to re-analyze the data. In total, we analyzed 7,216 individual cells derived from the small intestine of normal C57BL/6 mice. Using the unsupervised graph-based clustering, we found that the small intestine tissues consisted of at least eight distinct cell clusters according to their corresponding marker gene expression profiles (Figure 2A&B). For instance, the LGR5 gene was highly expressed in the stem cell cluster of the small intestine, and it was significantly reduced in other cell clusters (Figure 2B).

**Figure 2.**
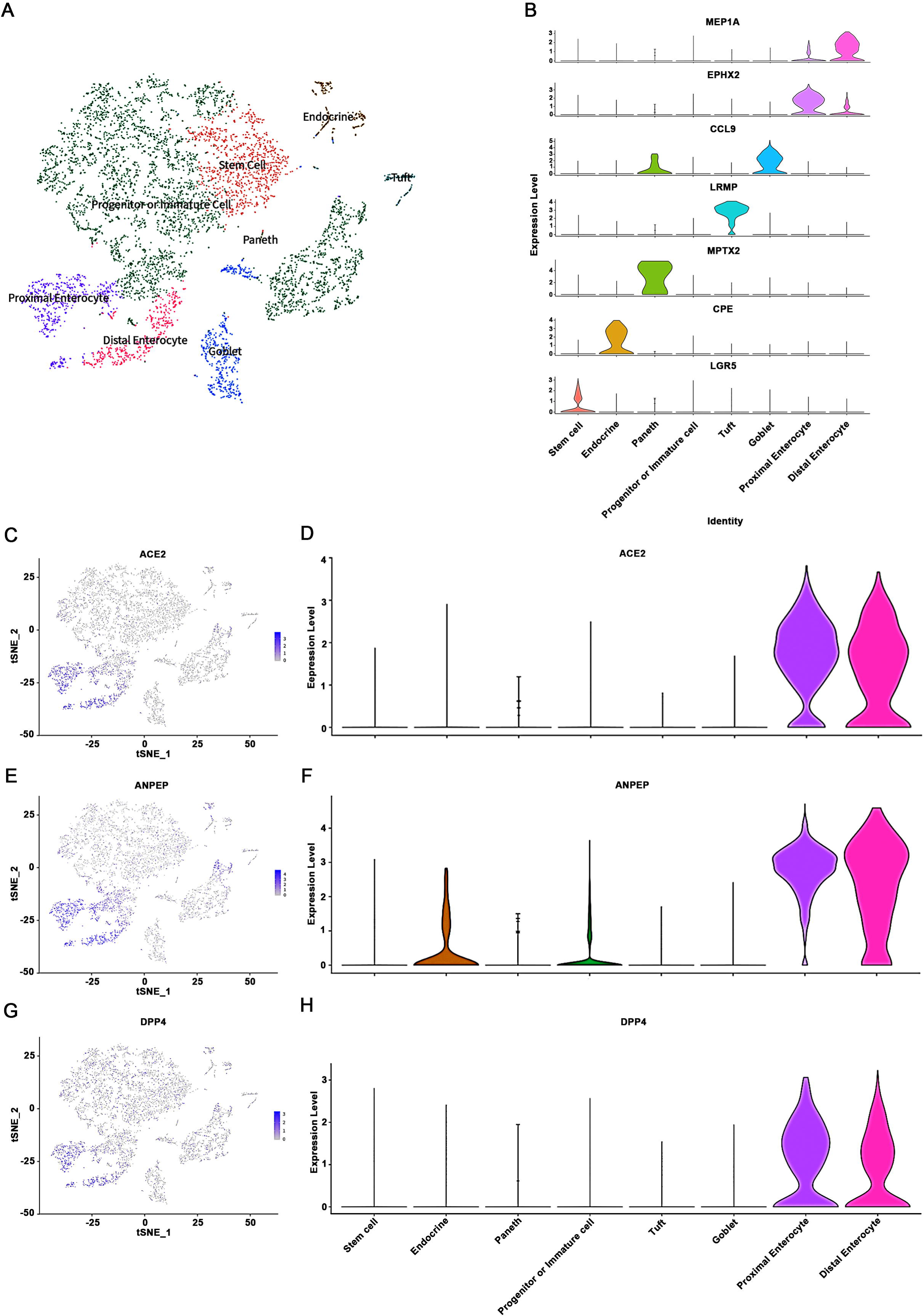
High expression of 2019-nCoV viral receptor ACE2 in enterocytes of small intestine. (A) Analysis of single-cell sequencing data identified eight cell sub-populations within mouse small intestine. (B) Cluster identity was identified by the expression of marker genes and the annotations provided in the UMI-barcode matrix. (C-H) Expression profile of three virus receptors ACE2 (2019-nCoV and SARS-CoV), ANPEP (HCoV-229E), and DPP4 (MERS-CoV) in eight subset cell clusters (C&E&G). Violin plots displayed that ACE2, ANPEP, and DPP4 are highly expressed in proximal and distal enterocytes (D&F&H).

We then focused on ACE2 and found that ACE2 was highly expressed in proximal and distal enterocytes (Figure 2C&D). Enterocytes, also called intestinal absorptive cells, are simple columnar epithelial cells located in the inner surface of the small and large intestines. Thus, enterocytes are directly exposed to food and foreign pathogens. Interestingly, when we examined expression profiles of another two virus receptors (ANPEP receptor for HCoV-229E virus and DPP4 receptor for MERS-CoV virus), we found that the RNA levels of these two virus entry receptors were also highly expressed in proximal or distal enterocytes (Figure 2E-H), consistent with the expression profile of ACE2. As these three virus receptors (ACE2, ANPEP, and DPP4) displayed significantly overlapped expression profiles, it is possible that the human intestinal tract may serve as an alternative infection route for these viruses.

## Discussion

Currently, the source and infection routes of 2019-nCoV remain unknown. The distribution of 2019-nCoV entry receptor determines the path of infection, and the route of infection is essential for understanding the pathogenesis, both of which are vital for infection control in hospitals and society. Based on the current findings, we proposed that: (1) The incidence of diarrhea may be underestimated in previous investigations, and it is an easily neglected symptom at an early stage; (2) ACE2-expressing small intestinal epithelium cells might be are more vulnerable to attack by 2019-nCoV.

In this study, we displayed that ACE2 was highly expressed in the small intestine, especially in proximal and distal enterocytes. Interestingly, another group has recently reported similar expression pattern in human small intestine one day before our submission ^13^. It has been characterized that enterocytes are directly exposed to food and foreign pathogens. Interestingly, other virus receptors like DPP4 displayed similar expression patterns as ACE2 in the small intestine. DPP4 is a known receptor for MERS-CoV through interacting with the S1 subunit of MERS-CoV spike protein. According to the recent publication, Zhou et al. reported the DPP4-expressing human intestine cells were highly susceptible to MERS-CoV and supported robust viral replication^14^, suggesting that the human intestinal tract may serve as an alternative infection route for MERS-CoV. In terms of the fact that most of the patients in the outbreak reported a link to a wild animal market, this observation raises an important question about whether this virus is transmitted via contaminated food when the food arrives at the small intestine. Therefore, further investigations are needed to address whether 2019-nCoV targets the human intestinal tract through ACE2 as MERS-CoV does via DPP4.

It has been reported that ACE2 controls intestinal inflammation and diarrhea. Therefore, mutual interaction between 2019-nCoV and ACE2 might disrupt the function of ACE2 and results in diarrhea or other gastrointestinal symptoms. According to the data analysis of three recent reports, we compared the incidence of various symptoms from infected patients and found that the incidence of diarrhea significantly differed in different reports. As 2019-nCoV is highly homologous to that of SARS-CoV, and it has been reported that around 20-25% of SARS patients have diarrhea^14^, it is confusing to observe the relatively low incidence (2-3%) of diarrhea in two cohorts from hospitals in Wuhan. If the incidence of diarrhea is underestimated, the reasons may because it is sometimes challenging to diagnose diarrhea since we still do not have a precise criterion for it. The definition of diarrhea by the World Health Organization is having three or more loose or liquid stools per day or having more stools than a person’s health condition. To a certain extent, this criterion is subjective.

Taken together, the symptoms of diarrhea could be underestimated. The information on discharge frequencies and the Bristol stool scale should be carefully collected. When infected patients with diarrhea go to the gastroenterology department, it will increase the risk of infection of healthcare workers. To reduce healthcare-associated infection, clinicians should be careful when their patients complain of diarrhea.

## Data Availability

The matrix we used for bioinformatic analysis can be loaded following the link https://www.ncbi.nlm.nih.gov/geo/query/acc.cgi?acc=GSE92332.

## Funding

This work was supported by grants from the National Natural Science Foundation of China (81870449 and 81670601 to Q.Z., 81902886 to W.L.), Special Fund for Frontier and Key Technology Innovation of Guangdong Province (2015B020226004 to Q.Z.), Key Project Fund of Guangdong Natural Science Foundation (2017A030311034 to Q.Z.), Guangdong Province Universities and Colleges Pearl River Scholar Funded Scheme

(Year 2017 to Q.Z.), National Key Point Research and Invention Program of the Thirteenth (2018ZX10723203 to Q.Z.).

## Ethics approval and consent to participate

Not applicable.

